# The influence of baseline clinical status and surgical strategy on early good to excellent result in spinal lumbar arthrodesis: a machine learning approach

**DOI:** 10.1101/2021.09.17.21263625

**Authors:** Francesco Langella, Luca Ventriglia, Domenico Compagnone, Paolo Barletta, David Huber, Francesca Mangili, Ginevra Licandro, Fabio Galbusera, Andrea Cina, Tito Bassani, Claudio Lamartina, Laura Scaramuzzo, Roberto Bassani, Marco Brayda-Bruno, Jorge Hugo Villafañe, Pedro Berjano, Laura Azzimonti

## Abstract

**Aims:** To create, using a machine learning (ML) approach, a preoperative model from baseline demographic and health-related quality of life scores (HRQOL) to predict a good to excellent early clinical outcome.

**Patients and Methods:** A single spine surgery center retrospective review of prospectively collected data from January 2016 to December 2020 from the institutional registry (SpineREG) was performed. The inclusion criteria were age ≥ 18 years, both sexes, lumbar arthrodesis procedure, a complete follow up assessment (ODI, SF-36 and COMI back) and the capability to read and understand the Italian language. A delta of improvement of the ODI higher than 12.7/100 was considered a “good early outcome”. A combined target model of ODI (Δ ≥ 12.7/100), SF-36 PCS (Δ ≥ 6/100) and COMI back (Δ ≥ 2.2/10) was considered an “excellent early outcome”. The performance of the ML models was evaluated in terms of sensitivity, i.e., True Positive Rate (TPR), specificity, i.e., True Negative Rate (TNR), accuracy and area under the receiver operating characteristic curve (AUC ROC).

**Results:** A total of 1243 patients were included in this study. The model for predicting ODI at 6 months follow up showed a good balance between sensitivity (74.3%) and specificity (79.4%), while providing a good accuracy (75.8%) with ROC AUC = 0.842. The combined target model showed a sensitivity of 74.2% and specificity of 71.8%, with an accuracy of 72.8%, and a ROC AUC = 0.808.

**Conclusion:** The results of our study suggest that a machine learning approach showed high performance in predicting early good to excellent clinical results.

## Introduction

Degenerative spine disorders represent a complex condition that mainly affects the elderly population, with an incidence in the healthy people over 70 years up to 68% [1]. Due to its clinical and socio-economic impact, it is playing an increasing role in daily medical practice. Its dissemination goes hand in hand with the aging of the population of developed countries. Spinal disorders have a broad spectrum of clinical manifestations: from minimal or asymptomatic to invalidating condition. The presentation pattern can variably affect segmental, regional, and global alignment. The pain and disability represent the main feature in a way that is comparable with other self-reported chronic conditions in the general population as congestive heart failure, arthritis, chronic lung disease or diabetes [2].

The therapeutic approach of spinal disorders is challenging in terms of decision-making for several causes and symptoms. Furthermore, the decisional process is made even more complicated by aging patients eligible for surgery and different clinical conditions and comorbidities. In the last decades, the rate of spine surgery increased up to 40%, and several randomized trials demonstrated the positive and significant effects of these procedures [3, 4]. Its safety and effectiveness vary widely among patients. In the worse scenarios, the complications rate can be up to 13%, and the minimally clinically relevant improvements can below up to 25% of cases [5]. Indeed, there is always room for improvements in terms of the clinical, surgical, and economic points of view [6].

The scientific research in this field targeted to evaluate the improvement in quality of life (QoL) after surgical treatment for spine surgery in relation to patient age, comorbidity and baseline status. With the aim to improve the cost-effect ratio’s performance, the rise of predictive models (PM) is continuously increasing. In 2015 McGirt et al. [7] presented a PM for the clinical practice to help patients, providers, and hospital systems. It’s based on demographics, patients’ reported outcomes, and clinical data. In particular, the baseline patient-specific factors such as symptom duration, smoking status, preoperative comorbidities and mental and physical conditions seem to significantly influence outcomes following lumbar surgery.

Sinikallio et al. [8], in a prospective analysis, demonstrated that the patients with preoperative depression and those who had continuous depression postoperatively experienced poor post-operative surgical outcomes and can benefit from targeted cognitive behavioural therapy [9]. Patient-specific factors beyond medical comorbidities, surgical indications, and surgical approaches can play a significant role in influencing overall patient outcomes [10]. The impact of lumbar spine surgery on patients’ life is commonly evaluated with three Patient-Reported Outcome Measures (PROMs): Oswestry Disability Index (ODI), the Physical Component Score of the Short Form of the Medical Outcomes Study (SF-36 PCS), and pain scales (VAS leg and back). The minimum clinically important difference (MCID) is commonly considered the threshold to measure the effect of the surgery for the single questionnaire.

The use of PROMs and their prediction through a machine learning approaches represent a milestone in the development of shared, informed, and individualized decision making potentially capable to support the surgeon to choose the right intervention, at the right time, for the right patient [7]. Our study aims to develop a preoperative machine learning (ML) model to predict a good to excellent early clinical outcome by using baseline demographic and health-related quality of life scores (HRQOL).

## Materials and Methods

### Clinical and demographic data and outcomes

The study was conducted in a single spine surgery center and it is based retrospective review of prospectively collected data from the institutional registry: SpineReg [11]. The inclusion criteria were age ≥ 18 years, both genders, lumbar arthrodesis procedure identified using ICD-9 code (8106, 8107 or 8108), a follow up assessment (ODI, SF-36 and Core Outcome Measures Index - COMI back) and the capability to read and understand the Italian language. A full set of peri-operative and post-operative data along with clinical outcomes from January 2016 to December 2020 were evaluated. Exclusion criteria were a weak degree of baseline disability or pain (ODI < 20/100 and COMI back < 3/10), number of levels fused not specified and subject not stratified according to Glassman classification.

The study protocol was conducted in accordance with the Helsinki Declaration of 1957 as revised in 2000. The procedures followed the ethical standards of the responsible committee on human experimentation and was approved by the ethics committee of Ospedale San Raffaele, Milan, Italy (protocol “SPINEREG”, approved on April 13^th^, 2016). The project was supported with funds from the Italian Ministry of Health (project code CO-2016-02364645). All patients gave their written informed consent for the participation in the registry. Baseline demographics, BMI, gender, comorbidities collected through Comorbidity Charlson Index (CCI) [12], diagnosis according to Glassman Classification [13], number of spinal levels of intervention, spinal level indexed surgery, clinical scores resulting from medical surveys, complication and revision surgeries were collected.

Table 1 shows that the rates of missing data ranges from 0.0% for the baseline scores of the PROMs and for the patient’s personal information, to 20.4% for Levels variables. Independent variables with at least one missing value were imputed using predictive mean matching for numerical variables, binary/multinomial logistic regression or ordered logit model for categorical variables. Outcome variables were not imputed to avoid the introduction of bias into the results. Only patients with the observed outcome variables were included in the analysis.

**TABLE 1.** Table with the features included in the analysis and associated percentage of missing values.

|  |  |  |  |  |  |  |
| --- | --- | --- | --- | --- | --- | --- |
| Glassman | Equipe | Age | Gender | BMI | ASA | ODIPre |
| 18.8% | 0.0% | 0.0% | 0.0% | 1.0% | 0.0% | 0.0% |
| COMIPre | SFPPre | SFMPre | Levels | From_Level | To_Level | Charlson |
| 0.0% | 0.0% | 0.0% | 20.3% | 20.4% | 20.4% | 19.0% |

The primary outcome was the early (six months post op) significant clinical improvement. In particular, value of improvement higher than 12.7 for ODI [14], 6 for SF36-PCS and 2.2 for COMI Back were considered as indicator of significant clinical improvement [15, 16].

To classify surgical operations results with a “good early outcome”, we defined a delta of improvement of the ODI higher than 12.7/100. On the other hand, to identify surgical operations with an “excellent early outcome” we used combined target that identifies an excellent clinical result when all the three underlying targets ODI (Δ ≥ 12.7/100), SF-36 PCS (Δ ≥ 6/100) and COMI back (Δ ≥ 2.2/10) showed a relevant improvement.

An exploratory analysis was performed. Patients were classified as having or not the binomial risk factor (Risk +) or (Risk -). Different scenarios were simulated to verify the performance of each method of calculation in three age categories. For each scenario, a 2 × 2 table was built (Good Outcome+/- vs. Risk +/-). Chi-squared test was used for statistical association between reaching the good outcomes and the presence of risk factor. The odds-ratios with their 95 % confidence intervals, and point estimations of the sensitivity and specificity of the alignment rules to discriminate patients with final good or poor clinical outcome, the positive (PPV) and negative (NPV) predictive values and positive and negative likelihood ratios (LR+, LR-) were calculated. Differences between preoperative and postoperative clinical outcomes were tested with the two-tailed Student’s t test for paired samples. The Mann-Whitney U-test was used in the cases of abnormally distributed variables. Normality was verified with the Kolmogorov–Smirnoff test. The threshold for statistical significance was set at p < 0.05 in all the tests.

### Machine learning approach

The available dataset is characterized by the non-negligible presence of missing values. Therefore, data imputation of independent variables is first performed to exploit all the instances and obtain more stable and reliable results [17]. In the dataset, different data types are available and each of them is treated with dedicated techniques. In the case of numerical variables, for each instance with at least a missing value a small subset of complete instances similar to the instance under investigation is selected. From this set, a randomly sampled instance is used to replace the missing values. Discrete variables are instead imputed with ad hoc models: logistic regression models for binary variables, multinomial logistic regression models for unordered categorical variables and ordered logit models (or proportional odds models) for categorical variables with ordered categories. The data imputation is implemented with “mice” R package [18].

A multivariate classification model is used to predict the target variables: 1) single ODI improvement or 2) combined ODI + SF36 PCS + COMI Back. For both the targets, we use a Random Forest (RF) classification method to predict the outcome of the surgical operation. RF is an ensemble model composed by multiple decision trees, each of them trained independently on randomly sampled subsets of variables. The single outputs of the multiple decision tree models are then combined with a majority vote to obtain the final decision of RF. This ensemble helps in improving the predictive performance of the individual decision tree models. Indeed, RF has been recognized as one of the best performing classifiers in extensive classification studies [19], and the R implementation provided by the “randomForest” package is empirically more accurate than other implementations [20]. We thus train a RF model using the default settings of the “randomForest” R package for both targets. To evaluate the most important features used by RF to classify instances, we used the Mean Decrease Gini index, which measures the contribution of each variable to the homogeneity of internal and leaf nodes of the tree.

We train RF in cross-validation (5 folds) and we select the classification threshold for each fold by optimizing the geometric mean of sensitivity and specificity in a nested cross-validation loop. The proposed nested cross-validation allows to robustly estimate the optimal classification threshold and assess RF performance, while balancing sensitivity and specificity. This is particularly relevant since both the target variables are slightly unbalanced (70.7% of the available data are associated with a good surgical outcome and 43.3% of the available data are associated with an excellent surgical outcome).

The performance of the model is evaluated in terms of sensitivity, i.e., True Positive Rate (TPR), specificity, i.e., True Negative Rate (TNR), accuracy and area under the receiver operating characteristic curve (AUC ROC). The exploratory and the further machine learning analysis were performed in R [21].

The entire study was performed according to TRIPOD guideline for the development of multivariate models for individual prognosis or diagnosis [22].

## Results

A total of 1243 patients undergone to lumbar arthrodesis surgery were included in this study. The 9.5% were disc pathologies, 38.4% were disc collapse, 32.8% were spondylolysis or spondylolisthesis, 7.6% degenerative scoliosis, 0.1% were facet pathologies, 11.1% non-union, 0.3% were cancer and 0.2% were infection. The rate of early good outcome was 70.7% (n=879). The 43.3% (n=538) of patient reached an “excellent” early outcome. The patients had a median age of 56 (interquartile range: 22) years and 771 (62.0%) were female. The mean baseline disability of study population was ODI 47.3 ± 17.1, the mean pain score was COMI back 7.7 ± 1.7 and the mean quality of life was SF- 36 PCS 32.7 ± 6.9 and SF-36 MCS 45.5 ± 11.8.

Since univariate exploratory analysis did not collect significant results, a multivariate classification models were used to identify both surgical operations with good and excellent early outcome, the results of these analyses are reported in the supplementary materials. The model for predicting ODI (good early outcome) at 6 months follow up (FU) makes use of the following features: classification of the patient’s clinical state (Glassman), equipe operating (Equipe), age, gender, Body Mass Index (BMI), ASA code (ASA), pre-operative medical PROMs: (ODI, COMI, SF- 36 Physical and SF-36 Mental), number of vertebrae stabilized during the operation (Levels), starting and ending point of the stabilized vertebrae (From_Level, To_Level) and comorbidity Charlson Index (Charlson).

The model showed a good balance between sensitivity (74.3%) and specificity (79.4%), while providing a good accuracy (75.8%) with ROC AUC = 0.842.

This combined target model (excellent early outcome) makes use of the same features used for the good early outcome model. The excellent early outcome model showed a sensitivity of 74.2% and specificity of 71.8%, with an accuracy of 72.8%, and a ROC AUC = 0.808. Furthermore, both the models for predicting good and excellent clinical outcomes show a good balance between sensitivity (74.3% for good and 74.2% for excellent outcome) and specificity (79.4% for good and 71.8% for excellent outcome), while providing a good accuracy (75.8% for good and 72.8% for excellent outcome). Details are reported in the Tables 2 and 3.

**Table 2.** 2 × 2 table showing the performance evaluation of AI model predicting the “good clinical outcome” – ODI at 6 months FU

|  | Outcome + | Outcome - | Total |  |  |
| --- | --- | --- | --- | --- | --- |
| Prediction + | **653** | **75** | 728 | Sensitivity | 74.3% |
| Prediction - | **226** | **289** | 515 | Specificity | 79.4% |
| Total | 879 | 364 | 1243 | PPV | 89.7% |
|  |  |  |  | PNV | 56.1% |
|  |  |  |  | Accuracy | 75.8% |
|  |  |  |  | AUC ROC | 0.842 |

Bold values indicate the number of patient with “Good Clinical Outcome” +/- and AI Prediction +/-.

**Table 3.** 2 × 2 table showing the performance evaluation of AI model predicting the “excellent clinical outcome” – ODI – SF36 – COMI Back at 6 months FU

|  | Outcome + | Outcome - | Total |  |  |
| --- | --- | --- | --- | --- | --- |
| Prediction + | **399** | **199** | 598 | Sensitivity | 74.2% |
| Prediction - | **139** | **506** | 645 | Specificity | 71.8% |
| Total | 538 | 705 | 1243 | PPV | 66.7% |
|  |  |  |  | PNV | 78.4% |
|  |  |  |  | Accuracy | 72.8% |
|  |  |  |  | AUC ROC | 0.808 |

Bold values indicate the number of patient with “Excellent Clinical Outcome” +/- and AI Prediction +/-.

The models show also a good discriminatory capacity of the two classes (ROC AUC = 0.842 for good and ROC AUC = 0.808 for excellent outcome) (Figure 1).

**Figure 1.**
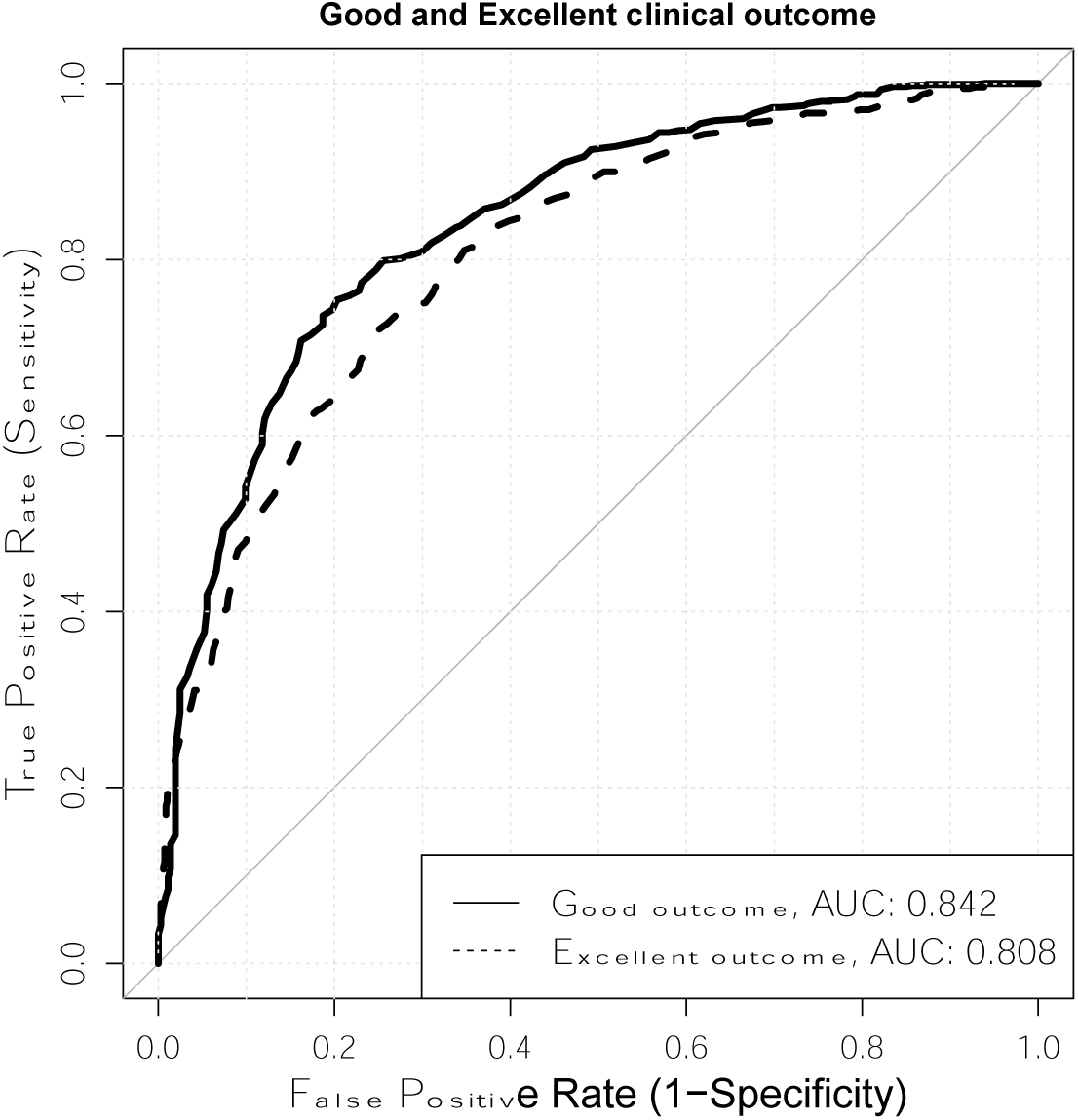
ROC CURVE For Model ODI and Combined Model.

According to the Mean Decreasing index of in the Random Forest model, the top five predictors of both good and excellent clinical outcomes were SF-36 PCS, SF-36 MCS, ODI, BMI, Age at baseline [weights of machine learning models for good clinical outcomes were: SFMPre = 73.20, SFPPre= 70.80, ODIPre = 66.77, BMI = 62.97 and Age =61.12; For excellent clinical outcomes were: SFMPre = 90.34, SFPPre= 87.13, BMI = 78.61, Age =69.92 and ODIPre = 66.21]. The tables and graphs of Mean Decrease Gini for weights of machine learning models, as well as odds ratios of the explorative analysis are resumed in the supplementary material.

## Discussion

In spine surgery, multiple factors can influence clinical outcomes. According to our results and the exploratory analysis, there isn’t a single risk factor capable of influencing or predicting early clinical outcomes. In our registry, the machine learning (ML) approach predicts the likelihood of good or excellent early clinical results. Our ML model showed good performances of post-operative prediction if based on patients’ demographic data and pre-operative self-reported degree of disability and quality of life.

In the last decades, the machine learning approach in predictive models has gained interest in clinical practice.

### 1. Predictive Models of surgical improvement based on clinical data

The surgical treatment for degenerative spine disorders has been shown to improve the quality of life and reduce disability in patients most severely affected [23]. Nevertheless, the association between demographic baseline factors and overall complication rates are still unclear.

Thanks to the anesthesiological and surgical implementations, a large population can safely be a spinal surgery candidate. The increase in the use of mini-invasive surgery [24] and the improvements of pre-operative planning methods [25] has allowed enlarging the cohort of patients eligible for surgery and capable of obtaining significant results [26, 27].

One of the most relevant demographic indicators is BMI (Body Mass Index). High BMI values are known to be risk factors for many diseases and directly correlated with the complication rate after spinal surgery. Even if in the literature, its role it’s still debated in the prediction of functional outcomes.

According to Mulvanay et al. [28], an increased BMI is associated with decreased effectiveness of 1- to 3-level elective lumbar fusion, despite the absence of surgical complications. A BMI value higher than 30 is considered a risk factor for surgical complications and poor spine surgery results. According to our data, a low BMI seems to present a relevant role in predicting good clinical outcomes. Despite, several studies suggest that weight does not represent a major impact on the patients’ health-related quality of life after surgery[29], obesity has a relevant impact on intraoperative blood loss, length of surgery and complication rate. It seems that BMI should always be kept in mind when planning spinal fusion.

Several clinical indicators of postoperative success are continuously analyzed to improve the surgical outcome in terms of complication rate and patients’ satisfaction. The aging of the population and the relative increase of the comorbidities can challenge the surgical decision.

According to Daniels et al. [30], the upgrading of some surgical methods increased the performances of therapeutic strategies. In particular, in a retrospective analysis of surgical cases enrolled between 2009 and 2016 the complication rates decreased over time, despite an increasingly elderly, medically compromised, and obese patient population. As a critical point, the authors identified the evolution of surgical strategies that resulted in an overall improvement of the treatment quality.

### 2. Predictive Models of surgical improvement based on PROMs

The proper estimation of the pre-operative degree of disability and quality of life is mandatory when surgery is required. In spine surgery, considerable controversy exists regarding spinal arthrodesis’ risk-benefit where surgery itself creates a permanent fusion of vertebral bodies. Nevertheless, several studies demonstrate a significant improvement after spinal arthrodesis in cases of degenerative spinal disorders [31–33]. A combination of scales is often used in clinical studies to assess multiple aspects of human being.

The indicators of quality of life and disability progressively gained attention, becoming the gold standard to measure the success rate after spine surgery. The post-operative clinical improvement can be evaluated based on patients’ reported outcomes such as ODI. Although post-operative improvement may be statistically significant, it is not necessarily clinically relevant. For this reason, several studies have defined the values to indicate a difference that is clinically meaningful to the patient (MCID) [34]. In particular, Monticone et. al defined as significant a cut off value of MCID at 12.7 ODI unit score of improvement [14].

### 3. Predictive models performances (General)

Predictive models for patient-reported outcomes can improve the surgical strategies when deciding to opt for surgery or not or potentially to adapt the surgical approach.

Despite the significant variability in the population affected by a common clinical condition, lumbar disc herniation, Staartjes et al. [35] proposed a predictive model based on deep learning-based analytics. Out of a population of 422 patients, the deep learning and logistic regression attained AUC values of 0.84 and 0.72, and an accuracy of 75% and 59%, respectively. The greatest discrepancy in performance measures regarded the models predicting back pain improvement. This could reflect the model’s weakness or the inherent difficulty of the outcome to be measured.

Models based on Naïve Bayes machine learning to predict hospitalization days and indications for discharge (for example, admission to rehabilitation facilities or back to home and hospitalization costs) showed high performances. In particular, the system proposed by Karnuta et al. revealed a predictive accuracy of 0.800 for costs of recovery, 0.874 for Length of Stay (LOS), and 0.878 for disposition with AUC for hospitalization costs (0.880), excellent AUC for LOS (0.941), and an excellent AUC for discharge disposition (0.906) [36].

The disease variability, combined with the psychological influencing factors and patients’ expectations related to the surgeries, challenges the accuracy of clinical predictions. Siccoli et al. [37] evaluated the feasibility of short- and long-term PROMs and reoperation rate by ML approach in patients affected by lumbar stenosis. According to this study, the models were able to predict the endpoints providing accurate information.

Although a progressive increase in the use of prediction models in spine surgery, little is known in spinal arthrodesis for 2 to 4 levels surgery.

Our results seem to provide comparable or higher predictive performances than other studies on spine surgery. Thanks to the recent advances in technologies, AI can involve the application of mathematical algorithms that continuously learn and make observations from existing data. The aim is to create a more accurate predictive model based on that data [38].

### 4. Influence of ML predictions on therapeutic strategy

With the widening of the modern dataset, the use of ML will progressively become the gold standard and the primary candidate for the data analysis. A shining future application in diagnosis, prognosis and decision-making process is desirable and will soon become an essential spine physician tool. Khan et al. introduced the application in the clinal management of cervical myelopathy and nontraumatic spinal cord injury to predict the risk of neurological impairment at one year [39]. These tools allowed the physicians to predict individual patient outcome after surgery for degenerative cervical myelopathy [40] and to apply preventive strategies as targeted physiotherapy and timing of psychological counselling.

With the application of ML techniques, several studies demonstrate the possibility to predict clinical outcomes. Ames C.P. et al. predicted the patients’ responses to SRS-22R (questionnaire) item per item up to 86.9% AUROC at 1 and 2 years following surgical treatment for ASD. The main clinical application is to aid surgical decision making during preoperative counselling [41]. In complex surgery, this approach will be capable of implementing already available surgical decision-making [42].

## Limitations

The result of our study comes with several limitations that we have to take into account. Firstly, the term follow-up indicators for 6 months can be considered only a preliminary result. External and prospective validations are necessary to support this methodology further to improve the knowledge acquired. Furthermore, the lower performance in terms of PPV in “excellent outcomes predictions” and PNV in “good outcomes predictions” can be explained by low number of positive and negative events respectively. Moreover, the two models can be combined to obtain a classification of patients in three categories: “Excellent”, “Good” and “Not good”. This classification of patients can be used to support clinicians in taking personalized and patient-specific decisions.

## Conclusion

The results of our study suggest that a machine learning approach showed high performance in predicting early good to excellent clinical results. In particular, our data suggest that a worse score of preoperative indicators of disability and quality of life, younger or healthier patients should aspect a significant clinically relevant improvement. On the other hand, older patients, higher BMI, comorbidities (higher ASA and Charlson score) with a higher score of SF-36 and lower score ODI would experience less clinically relevant improvement by forwarding the path of lumbar spine surgery. These results must be seen in the light of the study’s limitations, firstly, the middle term follow-up indicators, six months. A potential improvement or worsening in the PROMS results could occur later. The latter is not the focus of the study.

## Data Availability

Data are available from the corresponding author upon reasonable request.

## References

1. Schwab F, Dubey A, Gamez L, et al (2005) Adult scoliosis: prevalence, SF-36, and nutritional parameters in an elderly volunteer population. Spine (Phila. Pa. 1976).

2. Montse AV, Montse F, Bago J (2015) Impact on health related quality of life of adult spinal deformity (ASD) compared with other chronic conditions. 3–11. doi: 10.1007/s00586-014-3542-1

3. Weinstein JN, Lurie JD, Tosteson TD, et al (2009) Surgical compared with nonoperative treatment for lumbar degenerative spondylolisthesis. four-year results in the Spine Patient Outcomes Research Trial (SPORT) randomized and observational cohorts. J Bone Joint Surg Am 91:1295–304. doi: 10.2106/JBJS.H.00913

4. Dagenais S, Caro J, Haldeman S (2008) A systematic review of low back pain cost of illness studies in the United States and internationally. Spine J 8:8–20. doi: 10.1016/j.spinee.2007.10.005

5. Zanirato A, Damilano M, Formica M, et al (2018) Complications in adult spine deformity surgery: a systematic review of the recent literature with reporting of aggregated incidences. Eur Spine J 27:2272–2284. doi: 10.1007/s00586-018-5535-y

6. Ferguson TB (2012) The Institute of Medicine committee report “best care at lower cost: the path to continuously learning health care”. Circ Cardiovasc Qual Outcomes 5:e93–4. doi: 10.1161/CIRCOUTCOMES.112.968768

7. McGirt MJ, Sivaganesan A, Asher AL, Devin CJ (2015) Prediction model for outcome after low-back surgery: Individualized likelihood of complication, hospital readmission, return to work, and 12-month improvement in functional disability. Neurosurg Focus 39:1–10. doi: 10.3171/2015.8.FOCUS15338

8. Sinikallio S, Aalto T, Airaksinen O, et al (2011) Depression is associated with a poorer outcome of lumbar spinal stenosis surgery: a two-year prospective follow-up study. Spine (Phila Pa 1976) 36:677–82. doi: 10.1097/BRS.0b013e3181dcaf4a

9. Archer KR, Devin CJ, Vanston SW, et al (2016) Cognitive-Behavioral-Based Physical Therapy for Patients With Chronic Pain Undergoing Lumbar Spine Surgery: A Randomized Controlled Trial. J Pain 17:76–89. doi: 10.1016/j.jpain.2015.09.013

10. McGirt MJ, Bydon M, Archer KR, et al (2017) An analysis from the Quality Outcomes Database, Part 1. Disability, quality of life, and pain outcomes following lumbar spine surgery: Predicting likely individual patient outcomes for shared decision-making. J Neurosurg Spine 27:357–369. doi: 10.3171/2016.11.SPINE16526

11. Langella F, Barletta P, Baroncini A, et al (2021) The use of electronic PROMs provides same outcomes as paper version in a spine surgery registry. Results from a prospective cohort study. Eur Spine J. doi: 10.1007/s00586-021-06834-z

12. Charlson ME, Pompei P, Ales KL, MacKenzie CR (1987) A new method of classifying prognostic comorbidity in longitudinal studies: development and validation. J Chronic Dis 40:373–83. doi: 10.1016/0021-9681(87)90171-8

13. Glassman SD, Carreon LY, Anderson PA, Resnick DK (2011) A diagnostic classification for lumbar spine registry development. Spine J 11:1108–16. doi: 10.1016/j.spinee.2011.11.016

14. Monticone M, Baiardi P, Vanti C, et al (2012) Responsiveness of the Oswestry Disability Index and the Roland Morris Disability Questionnaire in Italian subjects with sub-acute and chronic low back pain. Eur Spine J 21:122–9. doi: 10.1007/s00586-011-1959-3

15. Copay AG, Glassman SD, Subach BR, et al (2008) Minimum clinically important difference in lumbar spine surgery patients: a choice of methods using the Oswestry Disability Index, Medical Outcomes Study questionnaire Short Form 36, and pain scales. Spine J 8:968–74. doi: 10.1016/j.spinee.2007.11.006

16. Mannion AF, Porchet F, Kleinstück FS, et al (2009) The quality of spine surgery from the patient’s perspective: part 2. Minimal clinically important difference for improvement and deterioration as measured with the Core Outcome Measures Index. Eur Spine J 18 Suppl 3:374–9. doi: 10.1007/s00586-009-0931-y

17. Harel O, Mitchell EM, Perkins NJ, et al (2018) Multiple Imputation for Incomplete Data in Epidemiologic Studies. Am J Epidemiol 187:576–584. doi: 10.1093/aje/kwx349

18. Buuren S van, Groothuis-Oudshoorn K (2011) mice : Multivariate Imputation by Chained Equations in R. J Stat Softw 45:. doi: 10.18637/jss.v045.i03

19. Fernández-Delgado M, Cernadas E, Barro S, Amorim D (2014) Do we need hundreds of classifiers to solve real world classification problems? J Mach Learn Res 15:3133–3181. doi: http://hdl.handle.net/10347/17792

20. Bagnall A, Cawley GC (2017) On the Use of Default Parameter Settings in the Empirical Evaluation of Classification Algorithms

21. Andy Bunn MK (2008) An Introduction to dplR. Ind Commer Train 10:11–18

22. Collins GS, Reitsma JB, Altman DG, Moons KGM (2015) Transparent reporting of a multivariable prediction model for individual prognosis or diagnosis (TRIPOD): the TRIPOD Statement. BMC Med 13:1. doi: 10.1186/s12916-014-0241-z

23. Reid DBC, Daniels AH, Ailon T, et al (2018) Frailty and Health-Related Quality of Life Improvement Following Adult Spinal Deformity Surgery. World Neurosurg 112:e548–e554. doi: 10.1016/j.wneu.2018.01.079

24. Berjano P, Lamartina C (2013) Far lateral approaches (XLIF) in adult scoliosis. Eur Spine J 22:242–253. doi: 10.1007/s00586-012-2426-5

25. Langella F, Villafañe JHJH, Damilano M, et al (2017) Predictive Accuracy of SurgimapTM Surgical Planning for Sagittal Imbalance: A Cohort Study. Spine (Phila Pa 1976). doi: 10.1097/BRS.0000000000002230

26. Berjano P, Ismael MF, Damilano M, et al (2014) Successful correction of sagittal imbalance can be calculated on the basis of pelvic incidence and age. Eur Spine J 23:S587–S596. doi: 10.1007/s00586-014-3556-8

27. Yamato Y, Md P, Hasegawa T, et al (2016) Calculation of the Target Lumbar Lordosis Angle for Restoring an Optimal Pelvic Tilt in Elderly Patients With Adult Spinal Deformity. Spine (Phila. Pa. 1976). 41:E211–E217

28. Mulvaney G, Rice OM, Rossi V, et al (2020) Mild and Severe Obesity Reduce the Effectiveness of Lumbar Fusions: 1-Year Patient-Reported Outcomes in 8171 Patients. Neurosurgery. doi: 10.1093/neuros/nyaa414

29. Lingutla KK, Pollock R, Benomran E, et al (2015) Outcome of lumbar spinal fusion surgery in obese patients: a systematic review and meta-analysis. Bone Joint J 97-B:1395–404. doi: 10.1302/0301-620X.97B10.35724

30. Tan GH, Goss BG, Thorpe PJ, Williams RP (2007) CT-based classification of long spinal allograft fusion. Eur Spine J 16:1875–81. doi: 10.1007/s00586-007-0376-0

31. Berjano P, Langella F, Ismael M-F, et al (2014) Successful correction of sagittal imbalance can be calculated on the basis of pelvic incidence and age. Eur Spine J 23 Suppl 6:587–96. doi: 10.1007/s00586-014-3556-8

32. Berjano P, Langella F, Damilano M, et al (2015) Fusion rate following extreme lateral lumbar interbody fusion. Eur Spine J 24 Suppl 3:369–71. doi: 10.1007/s00586-015-3929-7

33. Cecchinato R, Langella F, Bassani R, et al (2014) Variations of cervical lordosis and head alignment after pedicle subtraction osteotomy surgery for sagittal imbalance. Eur Spine J 23 Suppl 6:644–9. doi: 10.1007/s00586-014-3546-x

34. Carragee EJ, Cheng I (2010) Minimum acceptable outcomes after lumbar spinal fusion. Spine J 10:313–320. doi: 10.1016/j.spinee.2010.02.001

35. Staartjes VE, de Wispelaere MP, Vandertop WP, Schröder ML (2019) Deep learning-based preoperative predictive analytics for patient-reported outcomes following lumbar discectomy: feasibility of center-specific modeling. Spine J 19:853–861. doi: 10.1016/j.spinee.2018.11.009

36. Karnuta JM, Golubovsky JL, Haeberle HS, et al (2020) Can a machine learning model accurately predict patient resource utilization following lumbar spinal fusion? Spine J 20:329–336. doi: 10.1016/j.spinee.2019.10.007

37. Siccoli A, de Wispelaere MP, Schröder ML, Staartjes VE (2019) Machine learning-based preoperative predictive analytics for lumbar spinal stenosis. Neurosurg Focus 46:1–9. doi: 10.3171/2019.2.FOCUS18723

38. Lee MS, Grabowski MM, Habboub G, Mroz TE (2020) The Impact of Artificial Intelligence on Quality and Safety. Glob Spine J 10:99S–103S. doi: 10.1177/2192568219878133

39. Khan O, Badhiwala JH, Witiw CD, et al (2020) Machine learning algorithms for prediction of health-related quality-of-life after surgery for mild degenerative cervical myelopathy. Spine J 000:. doi: 10.1016/j.spinee.2020.02.003

40. Merali ZG, Witiw CD, Badhiwala JH, et al (2019) Using a machine learning approach to predict outcome after surgery for degenerative cervical myelopathy. PLoS One 14:1–12. doi: 10.1371/journal.pone.0215133

41. Ames CP, Smith JS, Pellisé F, et al (2019) Development of predictive models for all individual questions of SRS-22R after adult spinal deformity surgery: a step toward individualized medicine. Eur Spine J 28:1998–2011. doi: 10.1007/s00586-019-06079-x

42. Obeid I, Berjano P, Lamartina C, et al (2019) Classification of coronal imbalance in adult scoliosis and spine deformity: a treatment-oriented guideline. Eur Spine J 28:94–113. doi: 10.1007/s00586-018-5826-3

